# Reliability of auto-thresholds for remote surveillance of pacemakers: insights from the Calliope study

**DOI:** 10.1101/2025.09.15.25335484

**Authors:** Marie Wilkin, Marc Strik, Christoph Axthelm, Francisco Madeira, Ronald Hazeleger, Ignacio Fernández-Lozano, Sandra Rey Fariña, Romain Cassagneau, Simon Pearse, Antoine Guihard, Jean-Claude Deharo, Vincent Probst

## Abstract

In recent years, remote monitoring technology has transformed the management of CIEDs, by transferring part of historical in-hospital follow-up to dedicated teams and internet-based platforms allowing time saving.

Auto-threshold or auto-capture algorithms have been introduced in modern devices to improve patient management, particularly by enabling early detection of capture issues and dynamic output adjustment during fluctuations in pacing thresholds, by checking the minimal energy required to induce a depolarization wave in the RA or RV cavity. This allows permanently adapted pacing output, and potentially battery longevity extension if safety margins are reduced.

The MicroPort pacemakers (ALIZEA, BOREA and CELEA) provide remote monitoring via Bluetooth Low Energy, including RA and RV auto-threshold algorithms (RAAT and RVAT) and remote alerts.

The CALLIOPE investigation (NCT05165095) confirms the high accuracy of auto-threshold algorithms, in the range of 98%, which provide a reliable alternative to on-site follow-up in a vast majority of patients, when combined with the remote monitoring functions of ALIZEA, BOREA, and CELEA pacemakers.

## Background

In recent years, remote monitoring technology has transformed the management of CIEDs, by transferring part of historical in-hospital follow-up to dedicated teams and internet-based platforms allowing time saving.[1]

Auto-threshold or auto-capture algorithms have been introduced in modern devices to improve patient safety, particularly by enabling early detection of capture issues and dynamic output adjustment during fluctuations in pacing thresholds, by checking the minimal energy required to induce a depolarization wave in the RA or RV cavity. This allows permanently adapted pacing output, and potentially battery longevity extension if safety margins are reduced. [2-6]

The MicroPort pacemakers (ALIZEA, BOREA and CELEA) provide remote monitoring via Bluetooth Low Energy, including RA and RV auto-threshold algorithms (RAAT and RVAT) and remote alerts. The CALLIOPE investigation (NCT05165095) aimed to document the safety and performances of the remote monitoring functions of these devices.

## Methods and Results

CALLIOPE is a prospective international study with a total of 206 patients included across 18 sites in Europe. The patients were enrolled 0 to 23 days after dual-chamber pacemaker implantation and followed both on-site and remotely, with an on-site visit 1 to 3 months post-inclusion (M1-M3), resulting in an average follow-up of 2 months.

The mean age of the population was 76 ± 9 years. The indication for pacemaker implantation was third-degree atrioventricular block (38%), sinus node dysfunction (22%), high degree atrio-ventricular conductive disorder (22%), brady-tachy syndrome (9%) and other indications. Fifty percent of patients had ischemic cardiomyopathy.

The RAAT and RVAT functions can automatically adjust the amplitudes of atrial and ventricular pacing according to a periodical threshold test, performed every 24 hours for RAAT, and every 6 hours for RVAT. Both functions can operate either in “Auto mode” (RA and RV thresholds measurement result in automatic adjustment of pacing amplitudes), or in “Monitor mode” (no adjustment). Automatic adjustment in “Auto mode” consists in applying a programmable safety margin to the measured threshold; in case the threshold cannot be measured due to the absence of a suitable underlying rhythm, a safety amplitude (3.5V by default) is applied until the next retry.

RAAT and RVAT were activated after device implantation to allow remote monitoring of the RA and the RV pacing thresholds. The RAAT Auto mode was selected in 27% of the patients (RVAT Auto: 44%) while the Monitor mode was selected in remaining patients. Remote monitoring was activated in all patients, enabling device alerts per default (e.g., elevated pacing thresholds). The adequate transmission of device alerts was analyzed in order to report their sensitivity and specificity.

The accuracy of RAAT and RVAT was determined by the proportion of values showing a ± 0.5V difference compared to the manual measurements of pacing threshold during an on-site visit. Manual measurements of pacing threshold were performed at inclusion and at the M1-M3 visit by the investigator, and subsequently checked centrally by independent reviewers. The most recent automatic RAAT value stored in the device memory and the RV value resulting from an in-clinic launch of the RVAT function were used for analysis, which was based on all visits in which both a manual pacing threshold and an auto-threshold value were available. Each patient could contribute to two visits considering the pacing threshold is subject to variation between baseline and M1-M3 visits, resulting in a total of 309 visits evaluable from 179 patients for RAAT accuracy analysis, and 353 visits from 198 patients for RVAT accuracy.

Due to the different periodicity of assessment between RAAT and RVAT, the measurement rate was determined by the proportion of time with at least one automatic value found during follow-up (at least 1 value per week expected for RAAT, 1 value per day for RVAT). Periods of time in atrial fibrillation were excluded from RAAT analysis, as well as patients with incompatible pacing mode). A total of 179 and 194 patients contributed to this analysis for RAAT and RVAT, respectively.

The RAAT and RVAT accuracy obtained was 97.7% and 98.3% (Figure 1), while the measurement rate was 94.9% and 96.7%, respectively. Bland-Altman analyses confirm the absence of any clinically significant under or over estimation: no bias was found over the entire range of pacing thresholds (low Spearman correlation: 0.11 and 0.15, respectively). The average RA and RV pacing threshold during on-site assessments were 0.71 ± 0.27V and 0.78 ± 0.29V, respectively. For patients programmed in Auto mode, readjustments occurred in 61.8 % for RAAT and 44.4% for RVAT (the application of a safety amplitude was rather uncommon). The adequate transmission of technical remote alerts (device integrity and lead electrical parameters) was confirmed with a specificity of 100% and a sensitivity of 95.2%. Regarding alerts for elevated pacing threshold, 13 were raised by RAAT/RVAT and allowed to detect sustained elevation in seven cases, while the six others were triggered by single overestimated measurements.

**Figure 1:**
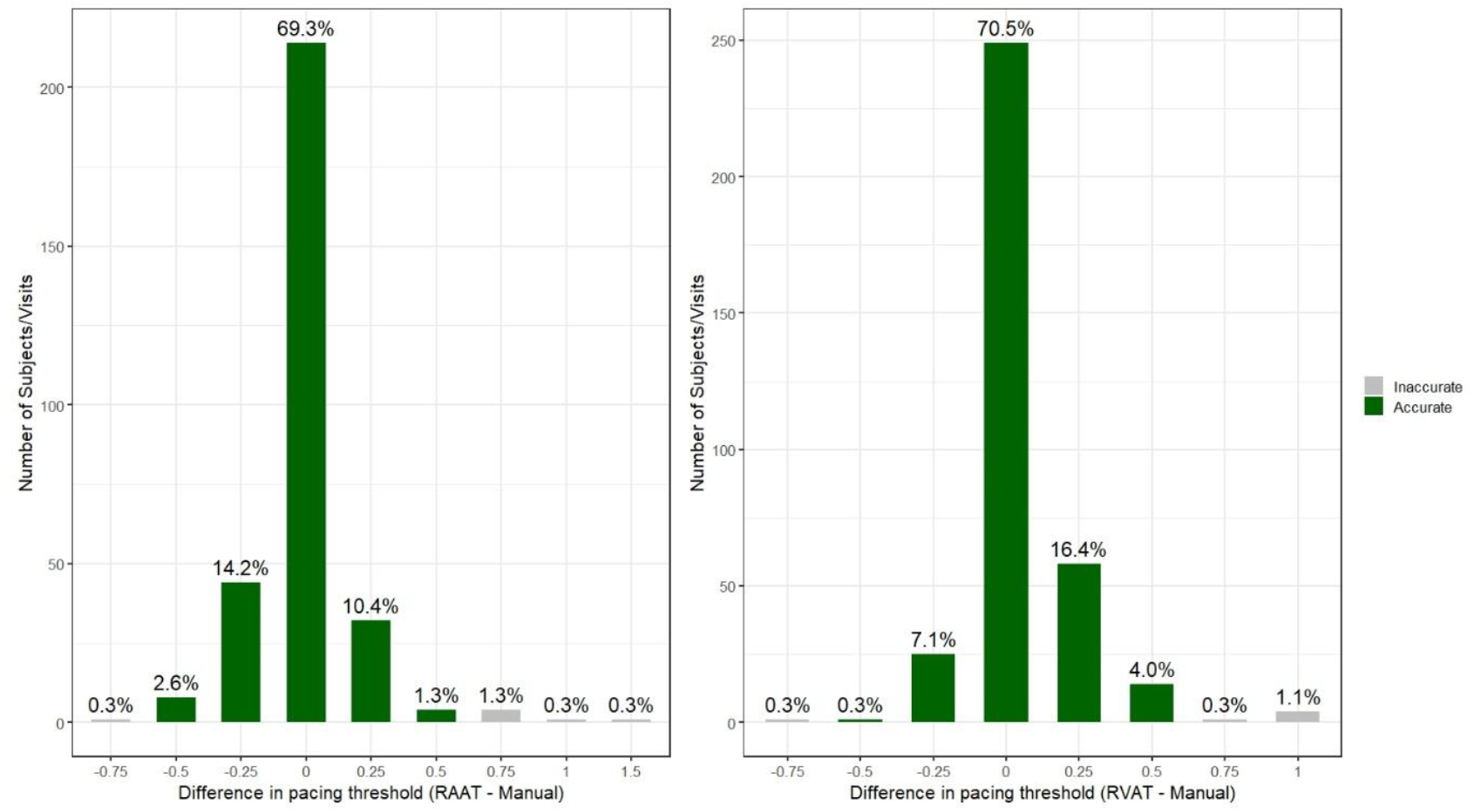
Distribution of pacing threshold differences (Autothreshold vs Manual); RAAT on the left, RVAT on the right.

## Discussion

The high accuracy of auto-threshold algorithms reported in the present study, in the range of 98%, compares well with previous studies investigating auto-threshold algorithms of pacemakers. [2-6] The main causes for inaccurate pacing threshold estimation by RAAT & RVAT involve competition with sinus rhythm, fusion beats or extrasystole onset during the test execution.

The transmission of remote alerts was found adequate, also in line with previous studies. [8] RAAT and RVAT performances remained stable over time, confirming that early performance adequately predicts future reliability. This allows the use of Auto or Monitor modes immediately after implant, and to check device memory at the next follow-up for informed decisions regarding the programming of auto-threshold, for optimization of energy consumption. Our study did not identify any predictor, including demographic data or lead model, that could determine RAAT/RVAT performance on an individual patient level.

Potential limitations of this study include the follow-up limited to two months, and the fact that each patient contributed to the analysis at two different follow-ups.

## Conclusion

Our study confirms that auto-threshold algorithms are accurate and provide a reliable alternative to on-site follow-up in a vast majority of patients, when combined with the remote monitoring functions of ALIZEA, BOREA, and CELEA pacemakers.

## Data Availability

All data produced in the present study are available upon reasonable request to the authors.

## Ethics Approval

The clinical investigation was reviewed and approved by the Ethics Committees (ECs). The list of the ECs that approved the study is provided below:

Approved by ethics committee of Austria: Ethikkommission der Medizinischen Universität Wien and Ethikkommission der Stadt Wien gemäß KAG, AMG und MPG; Belgium: Cliniques du Sud-Luxembourg Arlon (Comité d’éthique 246 OM 149) and Medical Ethics Committee of UZ Brussel/VUB; Germany: Ethik-Kommission der Ärztekammer Hamburg, Ethikkommission bei der Sächsischen Landesärztekammer and Ethikkommission an der Universitätsmedizin Rostock; Spain: Comité de ética de la investigación con medicamentos de Galicia; France: Comité de Protection des Personnes Nord Ouest IV; United Kingdom: London - Dulwich Research Ethics Committee and Health Research Authority; Netherlands: Medisch Ethische Toetsingscommissie van Isala in Zwolle; Portugal: Comissão de Ética para a Saúde do Hospital Professor Doutor Fernando Fonseca.

The clinical investigation was carried out in accordance with the ethical principles that have their origin in the Declaration of Helsinki, and that are consistent with the Good Clinical Practices (GCPs) described in the ISO 14155:2020, and the applicable regulatory requirements from the laws and regulations of the countries where the clinical investigation was conducted.

